# RESIST ACINETO test for the rapid detection of NDM and OXA acquired carbapenemases directly from blood culture in *Acinetobacter* species

**DOI:** 10.1101/2024.04.18.24302391

**Authors:** Anaïs Potron, Marion Daniel, Mila Bay, Pauline Choulet, Thomas Garrigos, Loïk Sababadichetty, Olivier Belmonte, Damien Fournier, Katy Jeannot, Guillaume Miltgen

## Abstract

The immunochromatographic assay RESIST ACINETO (Coris BioConcept) was evaluated on a collection of 121 *Acinetobacter* spp. clinical isolates, including 104 carbapenemase producers. The strains producing carbapenemases OXA-23, -40, -58, or/and NDM were accurately detected from bacterial cultures and directly from blood cultures, with the exception of an OXA-23/NDM-1-positive *A. radioresistens* isolate (only detected through standard culture). The performance of the RESIST ACINETO test was excellent (sensitivity 99%, specificity 100%) on this collection of isolates.

---

Carbapenem-resistant *Acinetobacter baumannii* (CRAB) poses a significant global threat, leading to a high mortality rate associated with antimicrobial resistance (1). This multidrug- or extensively drug-resistant pathogen is identified by the WHO as one of the three critical priority bacteria requiring urgent development of new antibiotics (2). In *Acinetobacter* species, the major mechanism for carbapenem resistance is attributed to the production of carbapenem-hydrolysing class D beta-lactamases, including OXA-23, OXA-24/40, and OXA-58 groups (3). Additionally, emerging NDM class B metallo-beta-lactamases play a significant role in conferring resistance (4,5). Since the activity of the last-line antibiotics, such as the newly developed cefiderocol, depend on the type of carbapenemase, rapid detection of these carbapenemase-producing bacteria has become crucial for optimizing antibiotherapy in case of infection and implementing appropriate hygiene measures (6). In recent years, several rapid immunochromatographic tests (Coris BioConcept, NG Biotech) have been developed for the detection of carbapenemase-producing Enterobacterales; but none has been specifically designed for non-fermenting Gram-negative bacilli, with the exception of the Coris OXA-23 K-SET test (accurate detection of OXA-23 group carbapenemases) (7–9). The novel RESIST ACINETO test (Coris BioConcept) has recently been developed to detect the main carbapenemases identified in the *Acinetobacter* species, namely the OXA-23-, OXA-24/40-, OXA-58- and NDM-groups (10). Here, we evaluated the performance of this novel immunochromatographic assay for the detection of carbapenemase-producing *Acinetobacter* spp. encountered in mainland and overseas France, from standard and blood cultures.

A large collection of 121 well-characterised *Acinetobacter* spp. clinical strains were selected from the French National Reference Center for Antibiotic Resistance (FNRC-AR) (*n*=61) and the University hospital of Reunion Island (*n*=60). The isolates were identified to the species level using matrix-assisted laser desorption ionization-time of flight (MALDI-TOF) mass spectrometry (MALDI Biotyper; Bruker Daltonics) or by sequencing. The collection included 107 isolates of *A. baumannii calcoaceticus* complex (*A. baumannii n*=95, *A. pittii n*=10, *A. nosocomialis n*=2), 7 *A. ursingii*, 1 *A. haemolyticus*, 1 *A. johnsonii*, 1 *A. junii*, 1 *A. proteolyticus*, 1 *A. radioresistens*, 1 *A. townerii* and 1 *A. variabilis*. One hundred and four strains produced one (*n*=75) or two carbapenemases (*n*=29) including 97 enzymes targeted by the test (27 OXA-23 type, 14 OXA-24/40 type, 6 OXA-58 type, 21 NDM type, 16 OXA-23+NDM-1, 3 OXA-58+NDM-1 and 10 others carbapenemase combinations) (Table S1). Three non-targeted carbapenemase, IMP-63 (*n*=1) et VIM-4 (*n*=2), were included, as well as 4 carbapenemase-hydrolyzing class D beta-lactamases (CHDL) rarely identified, OXA-235 (*n*=1), OXA-255 (*n*=2) and OXA-679 (*n*=1) (Table 1).

**Table 1.** Coris ACINETO results obtained with bacterial and blood cultures on the 121 isolates tested. P: positive; N: negative.

| Category | Species | Resistance mechanism | Number of strains | Coris ACINETO results obtained with bacterial culture |  |  | % Sensitivity (95% CI) | % Specificity (95% CI) | Coris ACINETO results obtained with blood culture |  |  | % Sensitivity (95% CI) | % Specificity (95% CI) |
| --- | --- | --- | --- | --- | --- | --- | --- | --- | --- | --- | --- | --- | --- |
|  |  |  |  | OXA-40/58 | OXA-23 | NDM |  |  | OXA-40/58 | OXA-23 | NDM |  |  |
| OXA-23 type (n=27) | *Acinetobacter* spp. | OXA-23 | 25 | N | P | N | 100 (88-100) | 100 (88-100) | N | P | N | 100 (88-100) | 100 (88-100) |
|  | *Acinetobacter* spp. | OXA-565 | 2 | N | P | N |  |  | N | P | N |  |  |
| OXA-24/40 type (n=14) | *Acinetobacter baumannii* | OXA-24 | 5 | P | N | N | 100 (78-100) | 100 (78-100) | P | N | N | 100 (78-100) | 100 (78-100) |
|  | *Acinetobacter baumannii* | OXA-72 | 9 | P | N | N |  |  | P | N | N |  |  |
| OXA-58 type (n=6) | *Acinetobacter baumannii* | OXA-58 | 3 | P | N | N | 100 (61-100) | 100 (61-100) | P | N | N | 100 (61-100) | 100 (61-100) |
|  | *Acinetobacter* spp. | OXA-420 | 3 | P | N | N |  |  | P | N | N |  |  |
| NDM-type (n=21) | *Acinetobacter* spp. | NDM-1 | 20 | N | N | P | 100 (84-100) | 100 (84-100) | N | N | P | 100 (84-100) | 100 (84-100) |
|  | *Acinetobacter baumannii* | NDM-9 | 1 | N | N | P |  |  | N | N | P |  |  |
| Multiples carbapenemase producers (n=29) | *Acinetobacter baumannii* | OXA-23 + OXA-58 | 3 | P | P | N |  |  | P | P | N |  |  |
|  | *Acinetobacter baumannii* | OXA-23 + OXA-420 | 3 | P | P | N |  |  | P | P | N |  |  |
|  | *Acinetobacter baumannii* | NDM-1 + OXA-23 | 15 | N | P | P |  |  | N | P | P |  |  |
|  | *Acinetobacter radioresistens* | NDM-1 + OXA-23 | 1 | N | P | P |  |  | N | N | N |  |  |
|  | *Acinetobacter baumannii* | NDM-1 + OXA-24 | 1 | P | N | P |  |  | P | N | P |  |  |
|  | *Acinetobacter* spp. | NDM-1 + OXA-58 | 3 | P | N | P |  |  | P | N | P |  |  |
|  | *Acinetobacter baumannii* | NDM-1 + OXA-420 | 1 | P | N | P |  |  | P | N | P |  |  |
|  | *Acinetobacter baumannii* | NDM-5 + OXA-23 | 1 | N | P | P |  |  | N | P | P |  |  |
|  | *Acinetobacter junii* | IMP-37 + OXA-58 | 1 | P | N | N |  |  | P | N | N |  |  |
| Other carbapenemase producers (n=7) | *Acinetobacter baumannii* | OXA-235 | 1 | N | N | N |  |  | N | N | N |  |  |
|  | *Acinetobacter pittii* | OXA-255 | 2 | P | N | N |  |  | P | N | N |  |  |
|  | *Acinetobacter pittii* | OXA-679 | 1 | P | N | N |  |  | P | N | N |  |  |
|  | *Acinetobacter baumannii* | IMP-63 | 1 | N | N | N |  |  | N | N | N |  |  |
|  | *Acinetobacter* spp. | VIM-4 | 2 | N | N | N |  |  | N | N | N |  |  |
| Non-carbapenemase producers (n=17) | *Acinetobacter baumannii* | Wild type | 1 | N | N | N |  |  | N | N | N |  |  |
|  | *Acinetobacter baumannii* | Overexpressed cephalosporinase | 5 | N | N | N |  |  | N | N | N |  |  |
|  | *Acinetobacter baumannii* | Overproduction of efflux pumps | 2 | N | N | N |  |  | N | N | N |  |  |
|  | *Acinetobacter baumannii* | CTX-M-15 + CARB-2 | 1 | N | N | N |  |  | N | N | N |  |  |
|  | *Acinetobacter baumannii* | GES-11 | 1 | N | N | N |  |  | N | N | N |  |  |
|  | *Acinetobacter baumannii* | PER-1 | 1 | N | N | N |  |  | N | N | N |  |  |
|  | *Acinetobacter baumannii* | CARB-5 | 2 | N | N | N |  |  | N | N | N |  |  |
|  | *Acinetobacter baumannii* | SHV-12 + CTX-M-15 | 1 | N | N | N |  |  | N | N | N |  |  |
|  | *Acinetobacter baumannii* | TEM-1 | 2 | N | N | N |  |  | N | N | N |  |  |
|  | *Acinetobacter johnsonii* | Wild type | 1 | N | N | N |  |  | N | N | N |  |  |

The performance of the RESIST ACINETO test was evaluated according to the manufacturer’s recommendations from standard and blood cultures. From bacterial colonies, a 1 μL full calibrated loop of bacteria grown for 16-24h at 35+/-2°C on Mueller-Hinton agar plate (bioMérieux) was added to 6 drops of lysis buffer. After homogenisation, 100 μl of the solution was added to the cassette. From blood culture, a blood culture vial (BACTEC Plus Aerobic/F, Becton Dickinson) was inoculated with 1 mL of a bacterial suspension calibrated at 10^2^ UFC/mL and incubated. Then, 1mL of the positive blood culture was collected and treated with the RESIST-BC protocol, as recommended by the manufacturer (Figure S1). Finally, 100 μL of the extraction solution was transferred into the cassette. The reading of cassettes was performed by two operators in a double-blind after 15 min. The sensitivity and specificity of the tests were calculated using the online program of the VassarStats website (http://vassarstats.net).

For the 97 carbapenemase-producing strains targeted by the test, all enzymes were correctly detected from standard and blood cultures, except for the *A. radioresistens* isolate (OXA-23 + NDM-1), where both enzymes were only detected from the standard culture (Table 1). However, carbapenem resistant *A. radioresistens* isolates are very rare, representing less than 0.1% of *Acinetobacter* species received at the FNRC-AR between 2013 and 2023. Seven NDM-producing strains have a low intensity band particularly from blood culture, with no inter-operator discordance and was considered positive according to the manufacturer’s recommendations (Figure 1). Interestingly, the test detected in both growth conditions, the strains producing the OXA-255 (*n=*2) and OXA-679 (*n=*1) enzymes, which share 87% and 73% amino acid identity with OXA-24/40, respectively (these strains were considered true positives for the performance calculation) (15). However, the test failed to detect the OXA-235 enzyme, which shares an amino acid identity lower than 65% with the enzymes OXA-24/40 (59%), OXA-23 (61%), and OXA-58 (64%). No false positives were observed for the strains producing a non-targeted carbapenemase (IMP-63 and VIM-4) or other beta-lactamases (CTX-M-, GES-, PER-, CARB-, RTG-, TEM-types) (Table 1). The test exhibited a sensitivity of 100% in standard cultures and 99% in blood cultures, while demonstrating a specificity of 100% in both conditions. Thus, the RESIST ACINETO test proves to be a reliable method for detecting carbapenemases in *Acinetobacter* species, as demonstrated by two recent studies (10,11) focusing on standard cultures. In our study, we extended the evaluation to blood culture, revealing consistent results with the previous findings.

**Figure 1.**
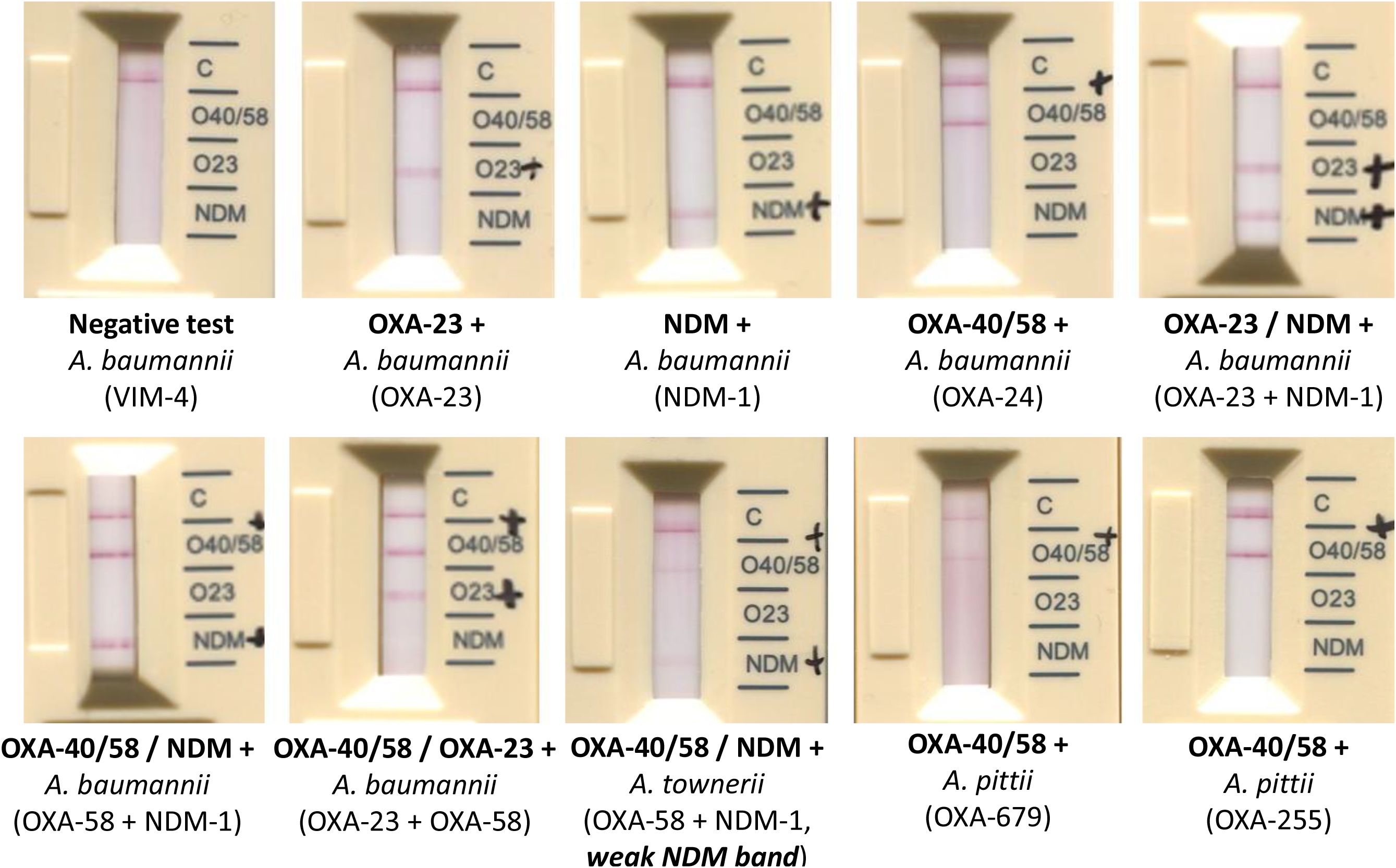
Representation of RESIST ACINETO test results for some isolates of interest.

These results opens up new potential applications for the test based on blood cultures (11). The use of the RESIST ACINETO test could prove highly beneficial for countries with a high frequency of carbapenem-resistant *Acinetobacter* species isolates, particularly in cases where the *A. baumannii-calcoaceticus complex* is responsible for a significant proportion of healthcare-associated bacteremia (*e*.*g*. South East Asia and the Balkans) (12,13). When performed directly after the presumptive identification of Gram-negative (coco-)bacilli in a positive blood culture, this test could facilitate the early detection imipenem-resistant isolates. It may also enable the optimization of treatment based on the type of carbapenemase present. It is well established that the MIC of cefiderocol against NDM-producing *A. baumannii* isolates are typically higher than 2 mg/L (6). This test could further assist in directing treatment towards alternative therapeutic options.

In conclusion, the RESIST ACINETO test is a rapid, easy-to-use, and cost-effective test that demonstrates excellent performance in detecting the major acquired carbapenemases present in the *Acinetobacter* species. It has the potential to be considered as an option for routine use in microbiology laboratories.

## Data Availability

All data produced in the present study are available upon reasonable request to the authors.

## Acknowledgments

This work was partially supported by the French Ministry of Health *via* Santé Publique France, and the ERDF European program (RESISTO-RUN project, 20201126-0022906).

We would also like to thank Coris BioConcept for supplying the kits. We precise Coris BioConcept was not involved in the design of the study or analysis of the results.

## Appendices

## Supplemental material

Figure S1: Blood culture sample processing protocol (adapted from RESIST-BC protocol, Coris BioConcept, technical note n°IFU-57S01/FR/V02, https://www.corisbio.com/products/resist-acineto).

Table S1: Molecular characteristics of the 121 *Acinetobacter* spp. isolates included in the study.

Strains originating in mainland France are marked as “France”, strains originating in overseas territories are marked with a distinctive sign (^¤^ Guadeloupe, ^♦^ Guyane, ^#^ Mayotte, * Reunion). The 4 light grey lines correspond to the rare oxacillinase variants (OXA-235, OXA-255, OXA-679) tested. The dark grey line corresponds to the *A. radioresistens* OXA-23+NDM-1 strain not detected in blood culture.

